# Cohort profile: The Swedish Evaluation Through Follow-up Study of Learning Later in Life (UGU-LIFE)

**DOI:** 10.64898/2026.06.24.26356445

**Authors:** Isabelle Hansson, Anne Ingeborg Berg, Pär Bjälkebring, Sandra Buratti, Jonas Burén, Linda Hassing, Anna-Carin Jonsson, Lina Jonsson, Magnus Lindwall, Andreas Segerberg, Valgeir Thorvaldsson, Mikael Landén, Alli Klapp, Martin Lövdé

## Abstract

**Purpose:** The Swedish Evaluation Through Follow-up study of Learning Later in Life (UGU-LIFE) was established to study the factors that shape lifelong learning and advance evidence-based means to facilitate learning in older age.

**Participants:** UGU-LIFE builds on the Evaluation Through Follow-up (*Utvärdering Genom Uppföljning*, UGU, in Swedish) study, which consists of nationally representative samples of Swedish birth cohorts. The two oldest cohorts, born 1948 (*N* = 11,945) and 1953 (*N* = 9,927), were assessed at age 13 years and invited for follow-up assessments as part of UGU-LIFE in 2025 (age 72/77 years; *N* = 5,738).

**Findings to date:** Data collection in childhood included a survey (on school and family conditions), cognitive tests, and school administrative data. The follow-up assessment in late adulthood included a survey (on personal and contextual factors), cognitive tests, learning tasks, and saliva sampling for DNA extraction. In addition, registry data was collected from Statistics Sweden (census), the National Board of Health and Welfare (medical records), and the Swedish National Archives (military conscription). Analysis of selectivity at follow-up showed higher retention rate among individuals with higher education and better cognitive ability in childhood, which was only partially explained by selectivity in survival.

**Future plans:** Data collected in UGU-LIFE will be used to describe the predictors of lifelong learning, the factors that influence learning gains and engagement in learning in older age, and the mechanistic pathways through which these factors affect learning in older age. Work to add birth records and geocoding to the data is ongoing. A subsample of participants will be invited to take part in an in-depth data collection of learning an ecologically relevant task over several days. The findings will be used to design and test interventions aiming to facilitate learning in older age.

**Strengths and limitations of this study:**

- UGU-LIFE includes population representative samples of individuals born in 1948 and 1953 (*N* = 21,872).
- Participants were followed over six decades to assess how factors in childhood predict learning in older age.
- Cognitive testing in both childhood and late adulthood offers unique insights into lifespan changes in cognitive functioning.
- DNA sampling allows for analysis of gene-environment interactions across the lifespan.
- Comprehensive registry data on health and socioeconomic conditions enable characterization of selective attrition, revealing both mortality-associated and experimental sources of selectivity at follow-up.

## Introduction

The knowledge and skills required for functioning in a society change during the life course due to both societal changes (e.g., technological advancements) [1] and individual life transitions (e.g., retirement) [2]. In adult life, acquiring new knowledge and skills is essential for personal and professional development. In older age, learning is fundamental for maintaining functional independence [3,4]. Much of this learning occurs as a part of daily life and is important, for example, in reasoning about medical decisions, identifying misinformation on social media, and avoiding online scams. Learning may even be required to perform mundane tasks such as buying bus tickets, ordering food in a restaurant application, accessing medical records, and booking appointments with healthcare providers.

Learning is a system’s processing of information from its environment, resulting in a change of system properties that alter its behavioral potential and reactions to this information [5]. It is distinct from performance but can only be observed by changes in behavioral performance, which give us the critical measure of learning gain [6]. Plasticity, the ability to learn, is theoretically separate from other cognitive abilities (e.g., working memory), but these other abilities may also influence learning gains [7,8,9].

Older adults face unique learning challenges, such as changing health, resources, cognitive functions, motivation, and sociocultural and individual expectations [10,11,12,13]. Differences among older adults are, however, best understood in a lifespan perspective, as the knowledge, skills, and expectations acquired in childhood set the stage for later learning [14,15,16,17]. While much focus has been put into understanding learning in the context of formal education in younger age [18,19,20], scientific knowledge about the unique challenges that older adults face in informal learning situations, the factors that explain individual differences among older adults in the ability to learn new skills, and the mechanisms that support learning later in life remain scarce. Societies are therefore generally unprepared for supporting their citizens’ lifelong learning. This lack of support negatively affects both societal and individual development.

The Evaluation Through Follow-up Study of Learning Later in Life (UGU-LIFE) was established to investigate the conditions shaping lifelong learning. By comprehensively charting the factors that explain individual differences in learning among older adults, the cohort provides a foundation for developing evidence-based means to facilitate learning throughout life, particularly for those who need it most. UGU-LIFE builds on the Evaluation Through Follow-up (*Utvärdering Genom Uppföljning*, UGU, in Swedish) study, a longitudinal research program designed to evaluate education and career development in Sweden [21]. UGU was initiated in 1961 by Statistics Sweden and the National Board of Education and includes nationally representative samples from 12 birth cohorts in Sweden.

The two oldest birth cohorts, born in 1948 and 1953, were included in UGU when they were 13 years old (in 6^th^ grade) in 1961 or 1966. In UGU-LIFE, the participants in these cohorts were reinvited for follow-up assessments in late adulthood. In this paper, we describe the baseline data collections in childhood and the first phase of follow-up assessments at age 72 (cohort 1953) or 77 (cohort 1948) years. This data collection establishes the empirical foundation for UGU-LIFE, enabling future planned studies to broaden the assessment of learning through complementary tasks, methodologies, and targeted interventions.

### Cohort description

The two UGU cohorts included in UGU-LIFE were sampled on birth date (born on the 5^th^, 15^th^, or 25^th^ of each month) to represent ten percent of the total Swedish population of each birth cohort (see Figure 1). The number of individuals born on these dates (i.e., sampling frame) was estimated to 12,166 in the 1948 cohort and 10,723 in the 1953 cohort. Data collection for the 1948 cohort was conducted between the 8^th^ and 27^th^ of May 1961. For the 1953 cohort, data collection took place between the 9^th^ and 28^th^ of May 1966. The data collections were administered by Statistics Sweden in collaboration with the Department of Education at the University of Gothenburg [21]. The students completed a survey and cognitive tests at school under the supervision of their teachers. School administrative data was collected by Statistics Sweden from year six until the end of compulsory school (year 9). A total of 11,950 individuals born in 1948 (98% of sampling frame) and 9,927 individuals born in 1953 (93% of sampling frame) took part in the study. However, for five individuals in the 1948 cohort, the data was not registry-linked, which reduced the dataset to 11,945.

**Figure 1.**
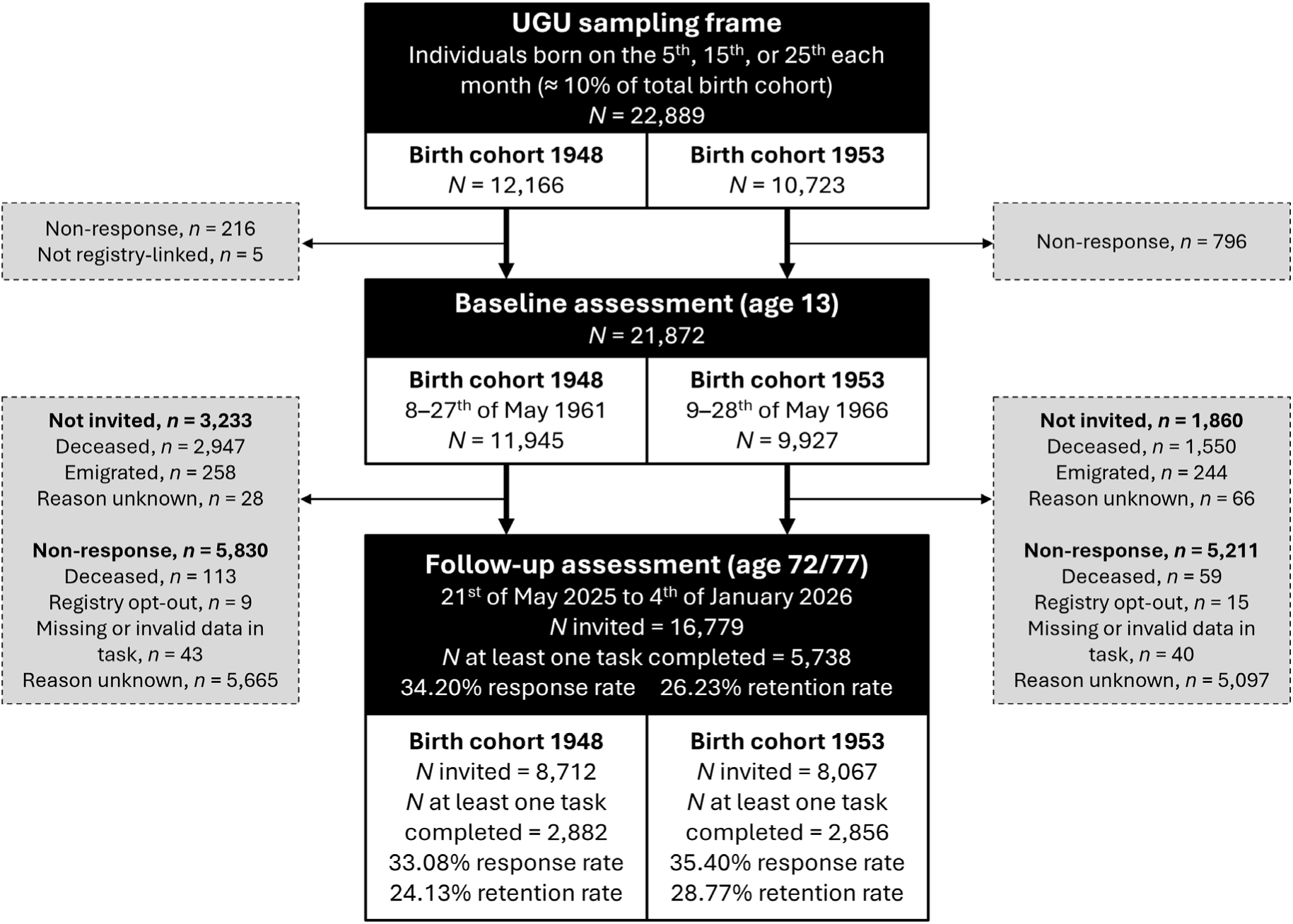
Flow chart of data collections in the Evaluation Through Follow-up Study of Learning Later in Life (UGU-LIFE). Registry linkage at follow-up was performed for all baseline respondents (N = 21,872), apart from 24 individuals requesting registry opt-out.

Six decades later, we invited the participants in the two cohorts for follow-up assessments as part of UGU-LIFE. The data collection took place between the 21^st^ of May 2025 and the 4^th^ of January 2026, when participants were 72 or 77 years old. In collaboration with Statistics Sweden, we identified individuals who were alive and living in Sweden by the end of March 2025 as eligible for follow-up (*N* = 16,779). Invitation letters, including a link to an online participation portal, were distributed via digital (if available; *n* = 8,889) and postal (*n* = 7,890) mail by Institutet för kvalitetsindikatorer (Indikator). Respondents completed a survey, cognitive tests, learning tasks, and indicated interest in donating saliva for DNA extraction. Registry data was collected from Statistics Sweden, the National Board of Health and Welfare, and the National Archives. Three reminders (June 16^th^, July 4^th^, and August 2^nd^) were sent via postal mail. The second reminder included a paper version of the survey, and the data collection portal was open until the 4^th^ of January 2026. Volunteers for saliva sampling received a separate invitation letter together with a self-administered sampling kit, distributed between the 23^rd^ of June and the 17^th^ of December 2025. Data collection for DNA extraction is ongoing and will be completed during fall 2026 (the current paper includes samples collected between the 27^th^ of June 2025 and the 12^th^ of May 2026).

A total of 5,738 (2,882 from the 1948 cohort and 2,856 from the 1953 cohort) individuals completed at least one task (out of seven) during the follow-up assessment, resulting in a retention rate of 26% and a response rate (relative invited) of 34%. The response rate was lower than generally observed in well-established national surveys (e.g., the SOM survey with a response rate around 50%) [22] but comparable to similar studies on older adults in Sweden [23]. It was also above the critical threshold of 30%, which has been found to substantially reduce nonresponse bias [24]. In the following sections, the data collections are described in more detail.

### Baseline childhood assessment (age 13 years)

The data collected as part of the childhood assessment consists of survey responses, cognitive test scores, and school administrative data via forms from Statistics Sweden. The questionnaire form filled in by the students in the 1948 cohort contained about 70 questions divided into four parts: leisure activities, plans, school, and interests. The leisure activities formed six scales covering outdoor, verbal, technical, domestic, social, and clerical areas. The school-related questions formed five scales: contact between children and parents, family’s attitude towards higher education, pupil’s anxiety in school situations, student’s interest in schoolwork, and student’s peer contacts. The questionnaire form filled in by the students in the 1953 cohort contained about 70 questions divided into four parts: leisure activities, plans, school, and occupations. The leisure activities formed five scales covering outdoor, verbal, technical, domestic, and clerical areas. The school-related questions formed three scales: family’s attitude towards higher education, pupil’s anxiety in school situations, and pupil adjustment to school.

The cognitive tests were the same for both cohorts and were designed to measure verbal knowledge (opposites), inductive reasoning (number series), and spatial ability (metal folding). The tests were of multiple-choice type with distractors and with a progression of level of difficulty. The opposite test consisted of 40 items of antonyms and students were asked to specify the opposite of a particular keyword among four options. The number series test contained 40 items and students were asked to complete a series of numbers, where six numbers were given, with two more numbers. The metal folding test included 40 items and students were asked to find out which of four figures you get if you fold a pictured flat and folded figures that match.

School administrative data have been gathered for both cohorts in yearly updates until the participants left school (year 9). These data include information on the participants concerning demographics and school and municipality characteristics, including school form, class type, class structure, class character, grade, school choice up through the education system, special tuition, special teaching group, reduced courses, moved from the class, and drop-out of compulsory school, mother’s and father’s education and occupation, birth month, and sex. Academic achievement outcomes, including standardized knowledge tests and subject grades, have also been collected. A detailed description of data collected as part of the baseline assessment is presented in Supplementary Material, Table 1.

### Follow-up assessment in late adulthood (age 72/77 years)

The data collected as part of the assessment in late adulthood consists of survey responses, cognitive test scores, learning performance, saliva sampling, and registry data on health and socioeconomic conditions. Figure 2 shows an overview of the number of respondents to each task and task combination (intersection). Figure 3 shows an overview of the lifespan data coverage in the two birth cohorts.

**Figure 2.**
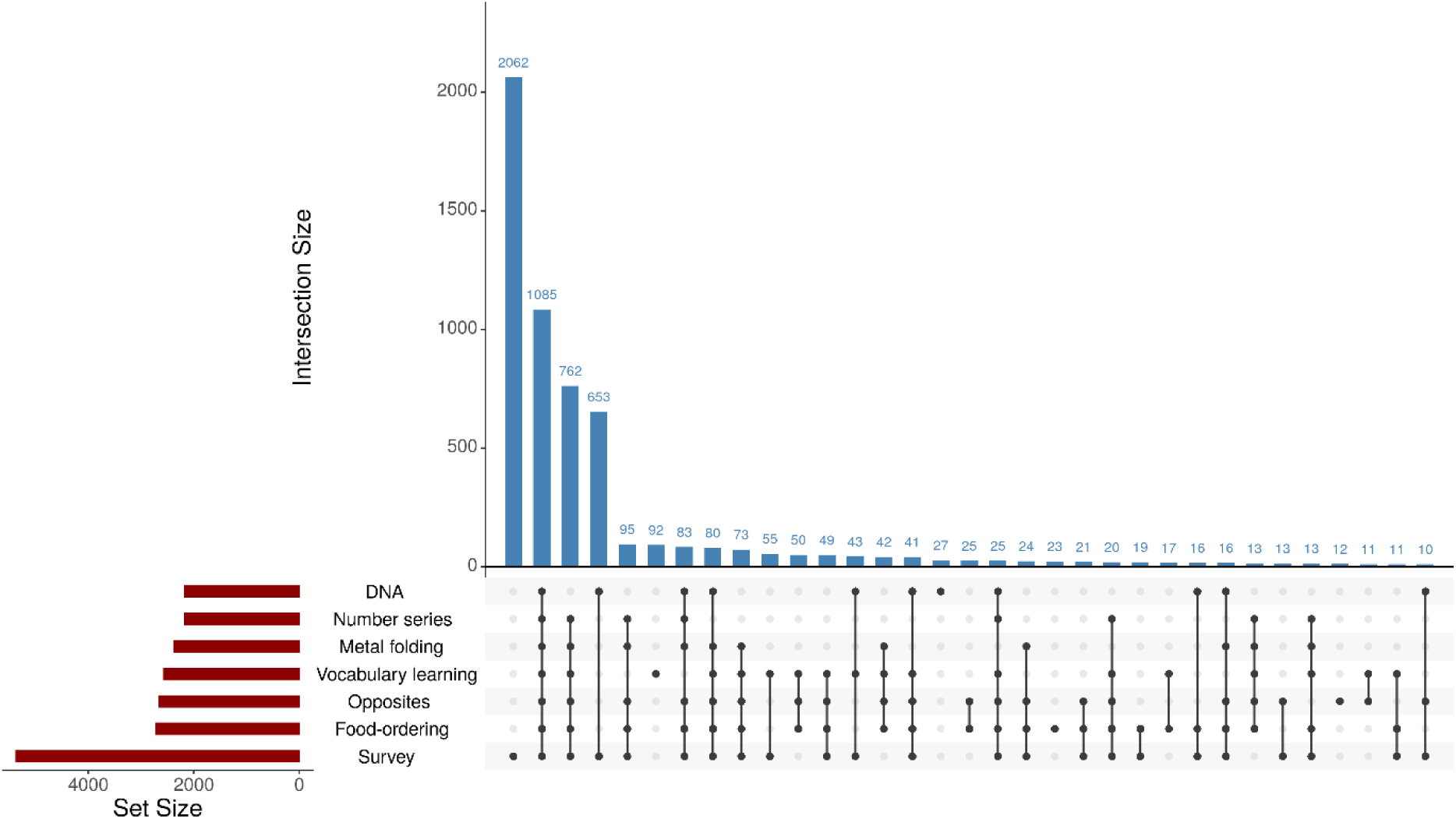
UpSet plot of participant frequency by task (Set Size) and task intersections (Intersection Size) in the follow-up assessment at age 72/77. For visualization purposes, only intersections (i.e., task combinations) with 10 or more observations (33 intersections) are included (in total 78 different task intersections were observed).

**Figure 3.**
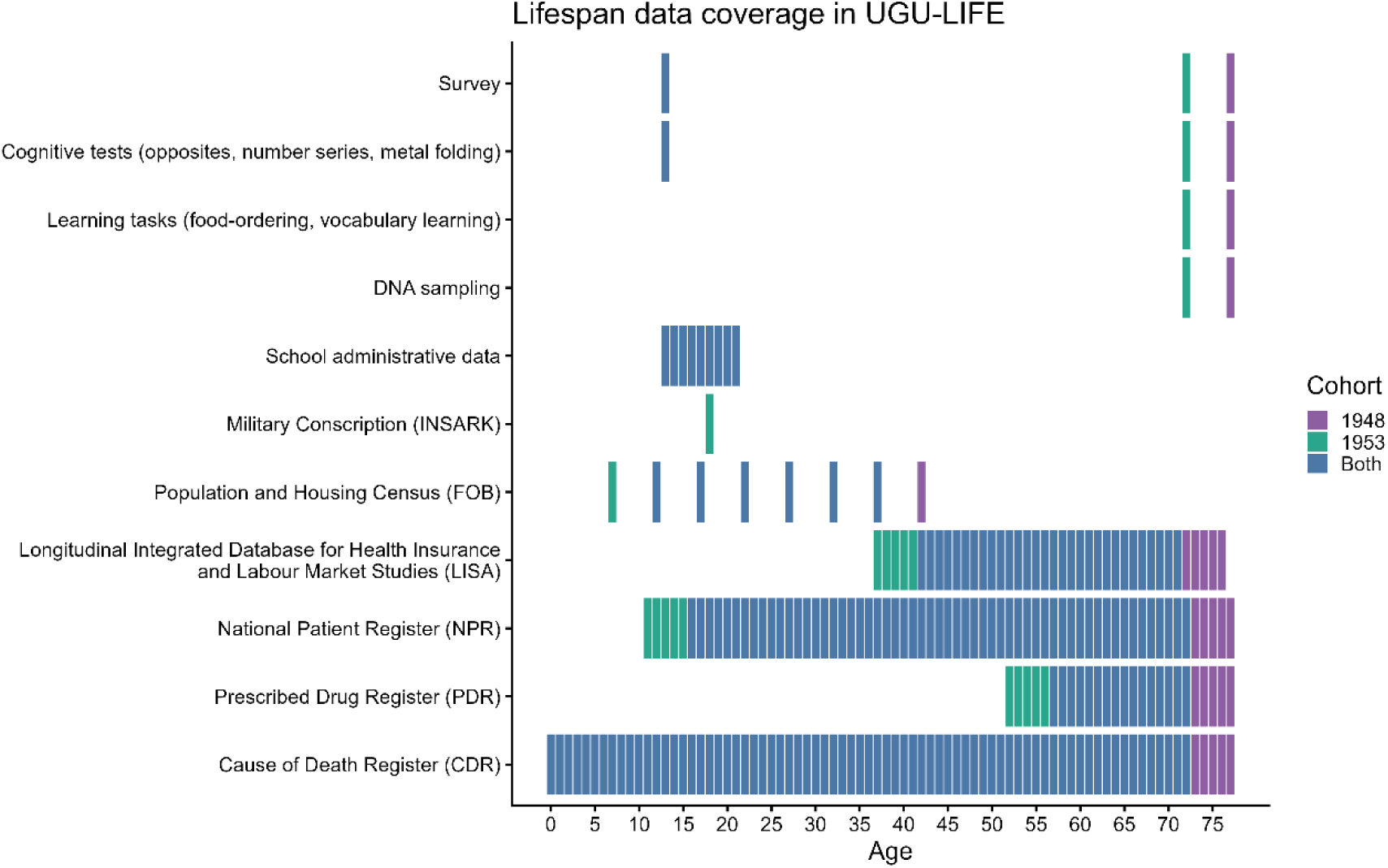
Overview of lifespan data coverage the Evaluation Through Follow-up Study of Learning Later in Life (UGU-LIFE). Survey and cognitive tests at age 13, and school administrative data part of original data collection in UGU. Survey and cognitive tests at age 72/77, learning tasks, DNA sampling, and linkage to registry data from Statistics Sweden (FOB, LISA), the National Board of Health and Welfare (NPR, PDR, CDR), and the Swedish National Archives (INSARK; men only) part of the follow-up assessment in UGU-LIFE. NPR has partial (regional) inpatient care from 1964 (age 11/16), full cover in psychiatric care from 1973 (age 20/25), and national coverage from 1987 (age 34/39). Outpatient care is available from 1997 (age 44/49) with full coverage from 2001 (age 48/53).

The questionnaire contained 56 questions with a total of 300 items. These were divided into seven parts: environmental context, learning engagement, motivation, expectations, personality and metacognition, resources, and health. Questions on environmental context covered family and household conditions, retrospective school assessments, leisure activities, and occupational history. Questions on learning engagement included frequency of learning engagement within theoretical (verbal, numeric, logical) and practical (mechanic, digital, motoric) domains, an adapted version of the Broad Learning Adult Questionnaire (BLAQ) [25], and internet use. Measures on motivation included the Achievement Motives Scale (AMS-R) [26] and the satisfaction subscale of the Basic Psychological Need Satisfaction and Frustration Scale (BPNSFS) [27]. Questions on expectations included perceived competence within theoretical (verbal, numeric, logical) and practical (mechanic, digital, motoric) learning domains, the Achievement Emotions Questionnaire (AEQ-S) [28], the Implicit Theories of Intelligence scale (ITI) [29], and the Awareness of Age-Related Change scale (AARC-10-SF) [30]. Personality was measured with a short form of the International Personality Item Pool-Five-Factor Model scale (MINI-IPIP) [31] and metacognition with a short version of the Metacognitive Awareness Inventory (MAI) [32]. Questions on resources included financial satisfaction, cash margin, the UCLA Loneliness Scale [33], the Multidimensional Scale of Perceived Social Support scale (MPSS) [34], and the Perceived Ageism Questionnaire (PAQ) [35]. Finally, questions within the health domain included the Satisfaction With Life Scale (SWLS) [36], the Patient Health Questionnaire for depression (PHQ-9) [37], and the Short Form Health survey instrument (SF-36) [38].

Cognitive performance was assessed using the same tests of verbal knowledge (opposites), inductive reasoning (number series), and spatial ability (metal folding) that had been administered in year 6 (age 13), although the follow-up assessments were conducted in an online format. To reduce the risk of ceiling effects, five additional items were added to the verbal knowledge test (at the end of the test). In addition, confidence judgements were added for 10 pre-selected items from each test. After the completion of each test, participants were presented with their answer to the selected items and asked to rate how sure they are that they have come to the correct answer (from 0 to 100%).

Learning performance was assessed with two short learning tasks, one primarily measuring procedural learning and one primarily measuring declarative learning. Procedural learning was measured using a food-ordering task designed as a hierarchical–sequential semantic decision task. The task simulated a food-ordering application (“UGU-Eats”) and required participants to navigate a structured menu to place specific orders. At the beginning of each trial, participants were presented with an item to be ordered as quickly as possible. To complete a trial, participants had to make eight correct decisions that followed a hierarchical menu structure. The task consisted of eight trials. Trials 1–4 involved the same learned sequence (ordering a big mackerel burger). Trial 5 introduced a novel sequence (medium raspberry sorbet), followed by the learned sequence in trial 6 (big mackerel burger). Trial 7 introduced another novel sequence (small apple juice), and trial 8 again used the learned sequence (big mackerel burger). The measure of performance for each trial was the total time (in seconds) required to complete the order, measured from trial onset to successful completion. Declarative learning was measured with a vocabulary learning task consisting of three study–test trials. The task involved learning associations between foreign words and Swedish translations. The foreign words were drawn from Sindarin, an artificial language created by J. R. R. Tolkien, to minimize prior familiarity. In each trial, participants were presented with a list of 10 Swedish–Sindarin word pairs. Following a study phase of 60 seconds, participants completed a forced-choice test. Each Sindarin word was presented individually together with 10 Swedish response alternatives. After the 10 responses had been provided, the next study–test trial began. Performance was measured through the number of correct answers per trial.

Saliva samples for DNA extraction were collected using a self-administered, non-invasive saliva collection kit (Oragene, DNA Genotek) that was mailed to participants. Samples were returned to the Karolinska Institutet Biobank (KIBB), where DNA was extracted continuously. Genotyping was performed using a genome-wide genotyping array (Illumina Infinium Global Screening Array – Multiple Disease version 3 [GSA-MD v3]) designed to capture common genetic variation across the genome.

Registry data was collected from Statistics Sweden, the National Board of Health and Welfare, and the Swedish National Archives for all participants in the original UGU samples (*N* = 21,872), except for individuals asking not to be included in registry linkage (opt-out offered as part of data collection; *n* = 24). Data from Statistics Sweden include information on demographic and socioeconomic conditions (incl. family background, residence, education, work life, income) from the Longitudinal Integrated Database for Health Insurance and Labour Market Studies (LISA; 1990–) and Population and Housing Census (FOB; 1960–1990). Data from the National Board of Health and Welfare include medical information from the National Patient Register (NPR; partial cover from 1964, national cover from 1987), the National Prescribed Drug Register (PDR; 2005–), and the National Cause of Death Register (CDR; full coverage). Finally, data from the National Archive include assessments of health and cognitive functioning from military conscription (INSARK; 1971). Registry data will be updated continuously as new waves of data become available via Statistics Sweden and the National Board of Health and Welfare. We also plan to collect birth record data from the regional archives. A detailed description of data collected as part of the follow-up assessment is presented in Supplementary Material, Table 2.

The whole data collection procedure (excl. saliva sampling and register linkage) was piloted on older adults from the same birth cohorts (i.e., 1948 and 1953) but born on other dates than the UGU participants (i.e., not on the 5th, 15th, or 25th of each month). Invitations were sent to 1000 individuals drawn from the population registry. To assess mode effects, half of the sample (*n* = 500) received paper-pencil versions of the cognitive tests. A total of 270 individuals completed the pilot study between the 31^st^ of January and the 28^th^ of April 2025. The responses were used to assess feasibility and psychometric properties of included measures, informing minor adjustments implemented in the main data collection.

### Findings to date

Data from UGU has been used to examine cohort differences in childhood cognitive ability [39], effects of educational reforms [40], and predictive effects of childhood conditions (e.g., family and school environment, cognitive ability and academic achievement) on educational [41,42] and health-related [43,44,45] outcomes. A complete list of UGU publications is available at https://www.gu.se/en/evaluation-through-follow-up/publications. However, no paper has yet been published on data collected as part of UGU-LIFE. The intent of this section is therefore to provide an overview of responses relating to the follow-up assessment in late adulthood.

The UGU-LIFE cohort (*N* = 21,872) includes 11,139 (51%) men and 10,733 (49%) women. In 2025, 16,779 individuals were identified as eligible (postal address in Sweden end of March) for follow-up, and 4,669 individuals were deceased (registered in CDR by October 26). Individuals born in 1948 had a higher mortality rate (26%) than those born in 1953 (16%). The response rate (relative invited) was somewhat higher in the 1953 cohort (35%) compared to the 1948 cohort (33%). The follow-up sample of 5,738 individuals includes 3,034 (53%) men and 2,704 (47%) women. The share of women at follow-up was slightly larger in the 1953 cohort (55%) than in the 1948 cohort (51%). A detailed description of sample characteristics at baseline and follow-up is presented in Supplementary Material, Table 3.

### Selectivity at follow-up

To assess selectivity at follow-up, we calculated standardized mean differences between the baseline sample and the follow-up sample (*M*_follow-up_ – *M*_baseline_ / *SD*_baseline_). This selectivity was further decomposed into a mortality-associated (*M*_survivors_ – *M*_baseline_ / *SD*_baseline_) and an experimental (*M*_follow-up_ – *M*_invited_ / *SD*_baseline_) component [46]. We thereby evaluate how much of the total selectivity that can be attributed to selectivity in survival (likelihood to be alive at follow-up) versus willingness to take part in the follow-up (experimental selectivity). Variables included in these analyses were: birth cohort, sex, birth region and region of residence (metropolitan vs. non-metropolitan), parental background (Swedish vs. foreign), parents’ education, childhood cognitive ability (total points of verbal, spatial and inductive tests), education, marital status, income, and health. Information on birth cohort, sex, parental education, and childhood cognitive ability were gathered from the original UGU data. Data on birth region, parental background, education, marital status and income come from Statistics Sweden. Health was calculated as weighted score of diagnoses (Charlson Comorbidity Index) [47] reported in the National Patient Register. For education, marital status, and health, we selected one indicator from early midlife (1990, age 37/42) and one from late adulthood (latest available data source, 2023–2025 depending on registry). Measures in early midlife thereby enable assessment of mortality risk (91% of deaths occurred after 1990) while measures in late adulthood capture how current conditions (among those still alive) shape experimental selectivity. Variable coding, descriptive statistics, and missingness patterns are presented in Supplementary Material, Table 3 and 4. Figure 4 shows an overview of the results, which are summarized below (all effect sizes are presented in Supplementary Material, Table 5). For interpretation of dichotomous variables, effect sizes are supplemented with rate ratios.

**Figure 4.**
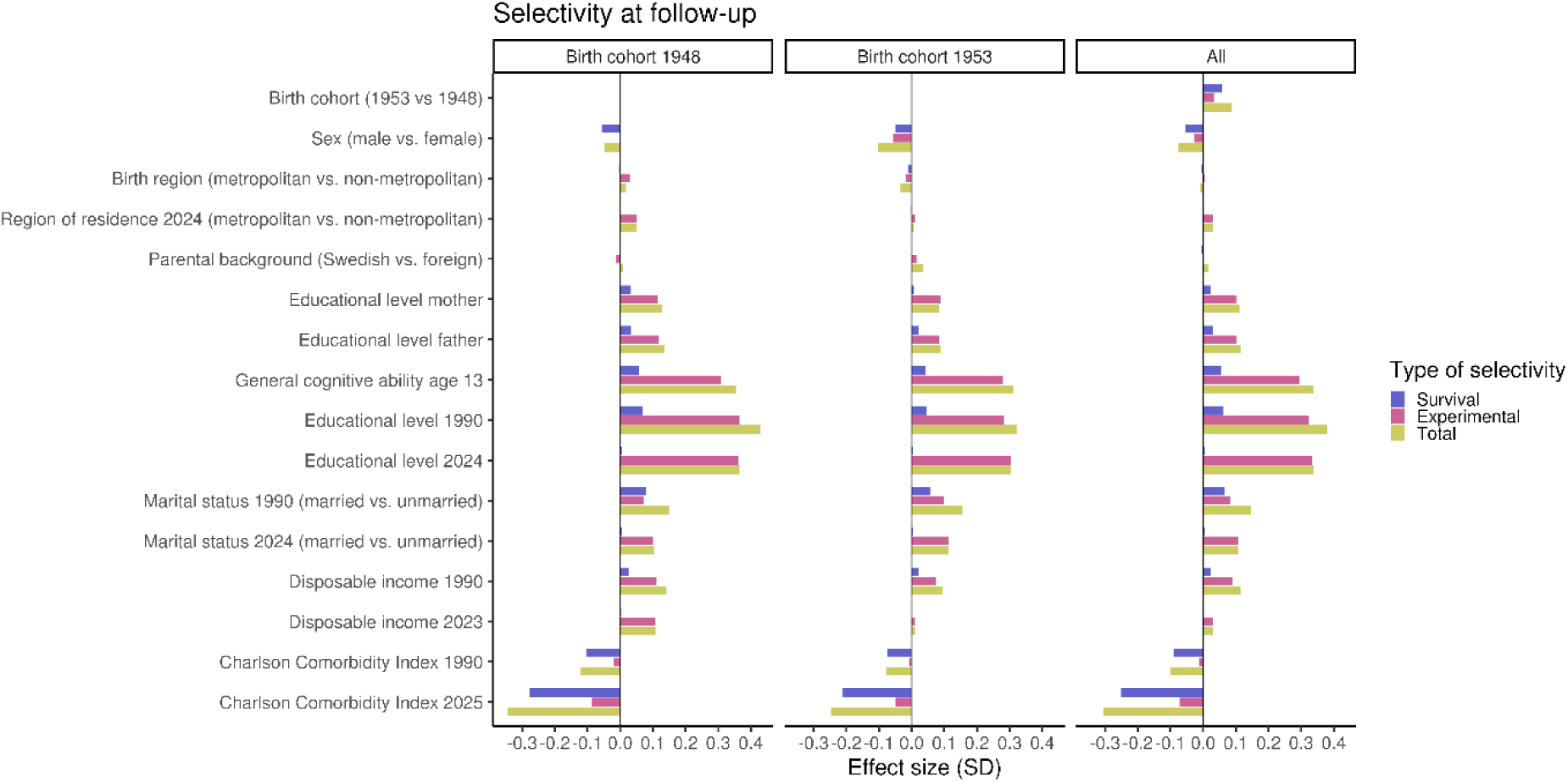
Selectivity (standardized mean difference) in survival (mortality-associated selectivity), follow-up (experimental selectivity), and retention (total selectivity) during the follow-up assessment at age 72/77 (2025).

Compared to those born in 1948, individuals born in 1953 were 13% more likely to be alive at follow-up and 7% more likely to complete the follow-up assessment (experimental selectivity). The mortality-associated component accounted for 68% of the total cohort-related selectivity at follow-up. Men were 10% less likely than women to be alive at follow-up and 5% less likely to complete the follow-up assessment. The mortality-associated component accounted for 71% of the total sex-related selectivity at follow-up.

Respondents at follow-up had higher parental education, higher childhood cognitive ability, and higher educational level (*SD* = 0.11–0.38). Only a smaller proportion (16–26%) of this selectivity could be attributed to selectivity in survival. Individuals married in 1990 were 39% more likely than unmarried to complete the follow-up assessment (total selectivity), and a large proportion (45%) of this selectivity could be attributed to selectivity in survival. Marital status in 2024 was also related to selectivity in follow-up, with married individuals being 25% more likely than unmarried to respond to the invitation. Higher income in 1990 was associated with a larger probability of taking part in the follow-up (*SD* = 0.11; mortality-associated component = 19%), but income in 2024 was only marginally related to response probability. Respondents at follow-up had better health in 1990 (*SD* = -0.10), but most of this selectivity (89%) could be attributed to selectivity in survival. Among individuals alive at follow-up, respondents had slightly better health compared to non-responders. Finally, only marginal selectivity effects were found for birth region, region of residence, and parental background.

### Selectivity in task completion at follow-up

Among the total follow-up sample of 5,738 individuals, 93% (*N* = 5,363) completed the survey (*N* = 2,207 online, *N* = 3,156 via paper), 47% (*N* = 2,711) the food-ordering task, 46% (*N* = 2,647) the opposites test, 45% (*N* = 2,564) the vocabulary learning task, 41% (*N* = 2,369) the metal folding test, 38% (*N* = 2,175) the number series test, and 38% (*N* = 2,166) donated saliva for DNA extraction. The average number of tasks completed per participant was 3.48 (*SD* = 2.50). Task-specific overlap is presented in Figure 2, cohort-specific frequencies are presented in Supplementary Material, Table 6.

Selectivity in task completion was assessed by calculating standardized mean differences between each task-specific sample and the total follow-up sample (*M*_task_ – *M*_follow-up_ / *SD*_follow-up_). Variables included in these analyses were: birth cohort, sex, region of residence, childhood cognitive ability, and the most recent registry data on education, marital status, income, and health. Figure 5 shows an overview of the results, which are summarized below (all effect sizes are presented in Supplementary Material, Table 7).

**Figure 5.**
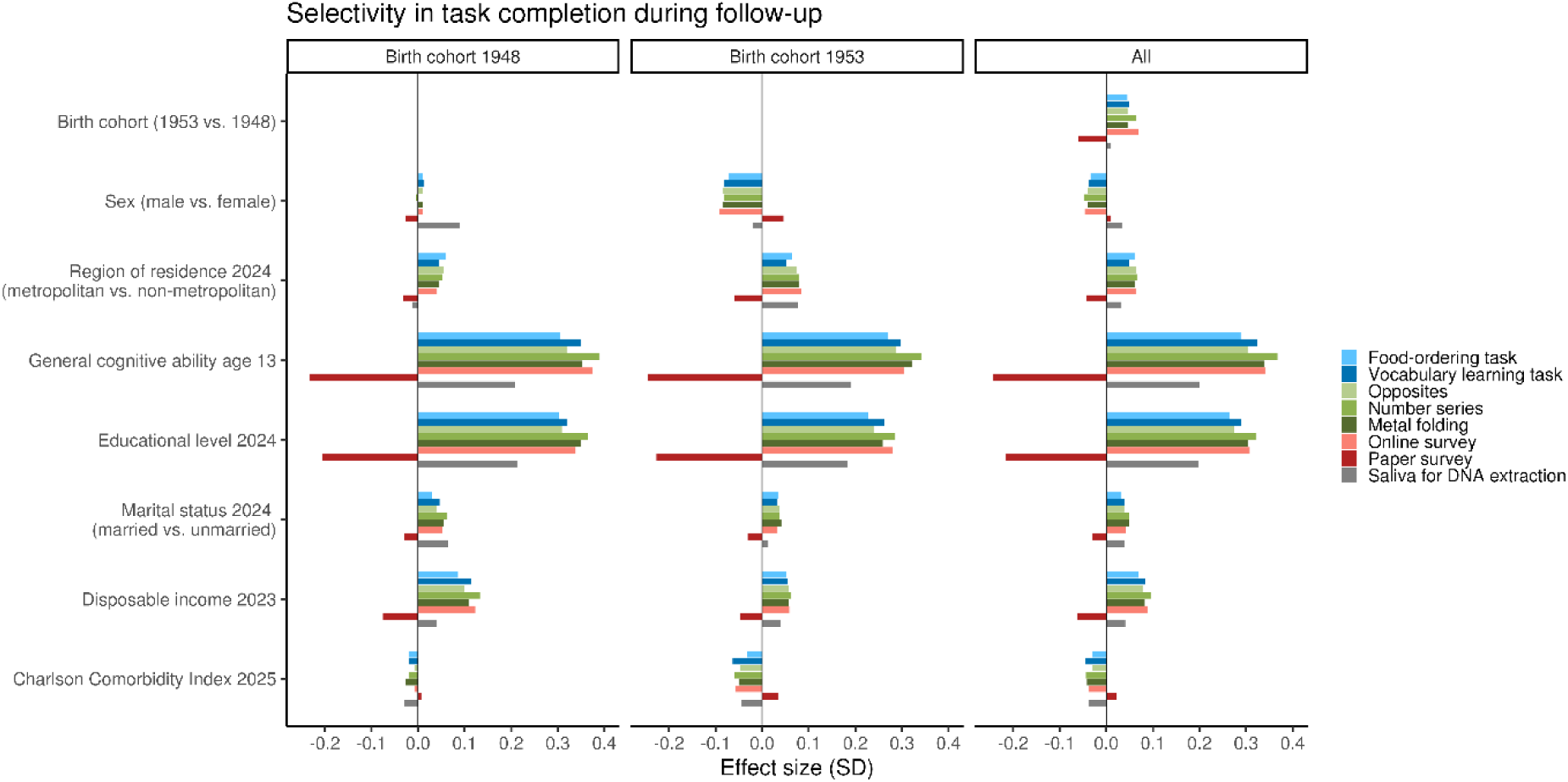
Selectivity (standardized mean difference) in task completion during the follow-up assessment at age 72/77 (2025).

Individuals born in 1953 were 10–15% more likely than those born in 1948 to complete online (learning tasks, cognitive tests, and online survey) tasks and 11% less likely to complete the paper survey, but no notable difference was found in the willingness to donate saliva for DNA extraction. Men were 7–9% less likely than women to complete online tasks and 7% more likely to donate saliva for DNA extraction, while almost no sex differences were found in paper survey completion. Sex differences were primarily shown in 1953 cohort, where men were 13–17% less likely to complete online tasks. For DNA sampling, the pattern was however reversed, with larger sex differences in the 1948 cohort (men 20% more likely to donate saliva).

Participants completing online tasks had better childhood cognitive ability and higher educational level compared to the total follow-up sample (*SD* = 0.26–0.37), an effect largely driven by poorer cognitive ability and lower education among paper survey respondents. Online responders also had higher income, slightly better health, and a larger proportion of married individuals and people residing in metropolitan areas, but the effect sizes were much weaker compared to those observed for cognitive ability and education. DNA donators had better cognitive ability and higher education (*SD* = 0.20), but only marginally higher income and better health compared to the total follow-up sample. Married individuals and people residing in metropolitan areas were slightly more likely to donate saliva.

### Strengths and limitations

UGU-LIFE is a rare population-based cohort that combines prospectively collected childhood data with a follow-up assessment six decades later, repeated cognitive testing across the lifespan, direct measures of learning in older age, genetic data, and comprehensive nationwide register linkage. The cohort builds on nationally representative samples of individuals born in Sweden in 1948 and 1953 and provides a unique opportunity to study how childhood conditions shape learning and related outcomes (e.g., health) in older age. The long follow-up period allows for investigation of developmental pathways across most of the life course. Another strength is the breadth of available data. Information from surveys, cognitive tests, learning tasks, DNA sampling, and nationwide registers can be combined within the same cohort. The register linkage provides detailed information on socioeconomic conditions, education, work life, health, medication, and mortality, enabling analyses of factors that may influence learning across adulthood. In addition, the cognitive tests administered in late adulthood are based on the same measures that participants completed in childhood, allowing assessment of long-term continuity and change in cognitive functioning.

UGU-LIFE was specifically designed to study learning in older age. In contrast to many aging studies that focus primarily on cognitive performance, the cohort data includes direct measures of learning performance together with assessments of learning engagement, motivation, expectations, metacognition, resources, and health. This combination provides opportunities to study both predictors of learning and mechanisms that may explain individual differences in learning gains among older adults.

The main limitation of UGU-LIFE is selective attrition. As expected after six decades, both mortality-associated and experimental sources of selectivity were observed. Individuals with higher education and cognitive ability were more likely to take part in the follow-up.

These patterns should be considered when interpreting findings based on the follow-up assessments. An important lesson is the value of offering multiple modes of participation when following older adults. Although completion of the online tasks was more common among individuals with higher education and cognitive ability, the option to respond using a paper questionnaire substantially increased participation and broadened coverage of the target population. Future studies aiming to assess learning and cognition in older age may benefit from combining digital and traditional modes of data collection.

### Collaboration

UGU-LIFE welcomes collaboration with researchers whose proposed work is of high quality and consistent with the scientific aims of the study, the ethical approvals governing the study, and applicable data protection legislation. The cohort data is particularly suited for research studying the factors influencing learning and related factors in later life from a lifespan perspective.

The UGU-LIFE dataset is currently being assembled, quality controlled, documented and integrated with registry and genetic data. Work to collect additional data on subsamples and work to retrieve birth records are ongoing. First analyses focused on validating questionnaires and tasks are currently conducted by the research group. In parallel, work on establishing the data sharing framework for the complete dataset is ongoing. Individual-level UGU-LIFE data are therefore not yet available to external researchers. We expect a controlled-access data sharing framework of the entire dataset to be established during the coming years as data collection is completed and data quality assurance, documentation, and governance procedures are finalized.

Once data access procedures have been finalized, researchers will be able to apply for access through a disclosure process administered by the University of Gothenburg (https://www.gu.se/en/evaluation-through-follow-up/disclosure-of-ugu-data). Data documentation and analytic codes will be published continuously via repositories on the Open Science Framework (https://osf.io/6mb3y) and GitHub (https://github.com/ugu-life).

## Further details

## Acknowledgements

Thanks to Lucas Fisher Madsen and Nelika Karimi for database management, and Kristine B. Walhovd for commenting on the manuscript.

## Contributor

Conceptualization: IH, AIB, PB, SB, JB, LH, ACJ, LJ, MLi, AS, VT, MLa, AK, MLö. Methodology: IH, AIB, PB, SB, JB, LH, ACJ, LJ, MLi, AS, VT, MLa, AK, MLö. Software: AS. Formal analysis: IH. Resources: AK. Data Curation: AS, IH, MLö. Writing - Original Draft: IH. Writing - Review & Editing: IH, AIB, PB, SB, JB, LH, ACJ, LJ, MLi, AS, VT, MLa, AK, MLö. Visualization: IH. Supervision: MLa, AK, MLö. Project administration: JB. Funding acquisition: MLö.

## Funding

The Evaluation Through Follow-up Study of Learning Later in Life (UGU-LIFE) has been funded by Riksbankens Jubileumsfond (Grant No. M23-0040).

## Competing interests

None declared.

## Patient and public involvement

Patients and/or the public were not involved in the design, or conduct, or reporting, or dissemination plans of this research.

## Patient consent for publication

Not applicable.

## Ethics approval

Ethical approval for UGU-LIFE was granted by the Swedish Ethical Review Authority (Dnr 2024-06028-01).

## Data availability statement

The UGU-LIFE dataset is not publicly available because it contains sensitive personal data that cannot be shared in the public domain under the conditions of the ethical approval and applicable data protection legislation. The UGU-LIFE dataset is currently being assembled, quality controlled, and documented. Work to collect additional data is ongoing. First analyses focused on validating questionnaires and tasks are currently conducted. In parallel, work on establishing data-sharing procedures is ongoing. Once the complete dataset is ready for release and the data-sharing procedures have been established, researchers will be able to apply for access to pseudonymised data through a controlled-access process administered by the University of Gothenburg (https://www.gu.se/en/evaluation-through-follow-up/disclosure-of-ugu-data), subject to scientific, ethical, legal and data protection requirements.

## Notes

### Competing Interest Statement

The authors have declared no competing interest.

## References

1. Charness N, Boot WR. Aging and information technology use: Potential and barriers. Curr Dir Psychol Sci. 2009;18(5):253–258. doi:10.1111/j.1467-8721.2009.01647.x

2. Wahl HW, Iwarsson S, Oswald F. Aging well and the environment: Toward an integrative model and research agenda for the future. Gerontologist. 2012;52(3):306–316. doi:10.1093/geront/gnr154

3. Rowe JW, Kahn RL. Successful aging. Gerontologist. 1997;37(4):433-440. doi:10.1093/geront/37.4.433

4. Nguyen C, Leanos S, Natsuaki MN, Rebok GW, Wu R. Adaptation for growth via learning new skills as a means to long-term functional independence in older adulthood: Insights from emerging adulthood. Gerontologist. 2020;60(1):4–11. doi:10.1093/geront/gny128

5. Lövdén M, Hansson I, Voits T. Learning: One definition to rule them all? npj Sci Learn. 2026.

6. Byrne JH. Learning and memory: A comprehensive reference. Amsterdam: Academic Press; 2017.

7. Ackerman PL. A theory of adult intellectual development: Process, personality, interests, and knowledge. Intelligence. 1996;22(2):227–257. doi:10.1016/S0160-2896(96)90016-1

8. Carroll JB. Human cognitive abilities: A survey of factor-analytic studies. Cambridge: Cambridge University Press; 1993.

9. Lövdén M, Bäckman L, Lindenberger U, Schaefer S, Schmiedek F. A theoretical framework for the study of adult cognitive plasticity. Psychol Bull. 2010;136(4):659–676. doi:10.1037/a0020080

10. Harris K, Krygsman S, Waschenko J, Laliberte Rudman D. Ageism and the older worker: A scoping review. Gerontologist. 2018;58(2):e1–e14. doi:10.1093/geront/gnw194

11. Kite ME, Stockdale GD, Whitley BE Jr, Johnson BT. Attitudes toward younger and older adults: An updated meta-analytic review. J Soc Issues. 2005;61(2):241–266. doi:10.1111/j.1540-4560.2005.00404.x

12. Rönnlund M, Nyberg L, Bäckman L, Nilsson LG. Stability, growth, and decline in adult life span development of declarative memory: cross-sectional and longitudinal data from a population-based study. Psychol Aging. 2005;20(1):3–18. doi:10.1037/0882-7974.20.1.3

13. Schaie KW. The course of adult intellectual development. Am Psychol. 1994;49(4):304–313. doi:10.1037/0003-066X.49.4.304

14. Fox PW, Hershberger SL, Bouchard TJ Jr. Genetic and environmental contributions to the acquisition of a motor skill. Nature. 1996;384(6607):356–358. doi:10.1038/384356a0

15. Sheffler P, Rodriguez TM, Cheung CS, Wu R. Cognitive and metacognitive, motivational, and resource considerations for learning new skills across the lifespan. Wiley Interdiscip Rev Cogn Sci. 2022;13(2):e1585. doi:10.1002/wcs.1585

16. Ullén F, Hambrick DZ, Mosing MA. Rethinking expertise: A multifactorial gene-environment interaction model of expert performance. Psychol Bull. 2016;142(4):427–446. doi:10.1037/bul0000033

17. Walhovd KB, Lövdén M, Fjell AM. Timing of lifespan influences on brain and cognition. Trends Cogn Sci. 2023;27(10):901–915. doi:10.1016/j.tics.2023.07.001

18. Blackwell LS, Trzesniewski KH, Dweck CS. Implicit theories of intelligence predict achievement across an adolescent transition: A longitudinal study and an intervention. Child Dev. 2007;78(1):246–263. doi:10.1111/j.1467-8624.2007.00995.x

19. Hill NE, Tyson DF. Parental involvement in middle school: a meta-analytic assessment of the strategies that promote achievement. Dev Psychol. 2009;45(3):740–763. doi:10.1037/a0015362

20. Alloway TP, Alloway RG. Investigating the predictive roles of working memory and IQ in academic attainment. J Exp Child Psychol. 2010;106(1):20–29. doi:10.1016/j.jecp.2009.11.003

21. Härnqvist K. Evaluation through follow-up. A longitudinal program for studying education and career development. Gothenburg: Department of Education and Educational Research, Gothenburg University; 1998. http://hdl.handle.net/2077/26970

22. Lundmark S, Backström K. Predicting survey nonresponse with registry data in Sweden between 1993 and 2023: Cohort replacement or a deteriorating survey climate?. Surv Res Methods. 2025;19(3):247–265. doi:10.18148/srm/2025.v19i3.8278

23. Lindwall M, Berg AI, Bjälkebring P, Buratti S, Hansson I, Hassing L, et al. Psychological health in the retirement transition: Rationale and first findings in the Health, Ageing and Retirement Transitions in Sweden (HEARTS) study. Front Psychol. 2017;8:1634. doi:10.3389/fpsyg.2017.01634

24. Hedlin D. Is there a safe area where the nonresponse rate has only a modest effect on bias despite non-ignorable nonresponse? Int Stat Rev. 2020;88(3):642–657. doi:10.1111/insr.12359

25. Leanos S, Coons J, Rebok GW, Ozer DJ, Wu R. Development of the broad learning adult questionnaire. Int J Aging Hum Dev. 2019;88(3):286–311. doi:10.1177/0091415018784695

26. Lang JW, Fries S. A revised 10-item version of the Achievement Motives Scale. Eur J Psychol Assess. 2006;22(3):216–224. doi:10.1027/1015-5759.22.3.216

27. Chen B, Vansteenkiste M, Beyers W, Boone L, Deci EL, Van der Kaap-Deeder J, et al. Basic psychological need satisfaction, need frustration, and need strength across four cultures. Motiv Emot. 2015;39:216–236. doi:10.1007/s11031-014-9450-1

28. Bieleke M, Gogol K, Goetz T, Daniels L, Pekrun R. The AEQ-S: A short version of the Achievement Emotions Questionnaire. Contemp Educ Psychol. 2021;65:101940. doi:10.1016/j.cedpsych.2020.101940

29. Dweck CS. Self-theories: Their role in motivation, personality, and development. Philadelphia: Psychology Press; 1999.

30. Kaspar R, Gabrian M, Brothers A, Wahl HW, Diehl M. Measuring awareness of age-related change: Development of a 10-item short form for use in large-scale surveys. Gerontologist. 2019;59(3):e130–e140. doi:10.1093/geront/gnx213

31. Donnellan MB, Oswald FL, Baird BM, Lucas RE. The mini-IPIP scales: Tiny-yet-effective measures of the Big Five factors of personality. Psychol Assess. 2006;18(2):192–203. doi:10.1037/1040-3590.18.2.192

32. Schraw G, Dennison RS. Assessing metacognitive awareness. Contemp Educ Psychol. 1994;19(4):460-475. doi:10.1006/ceps.1994.1033

33. Hughes ME, Waite LJ, Hawkley LC, Cacioppo JT. A short scale for measuring loneliness in large surveys: Results from two population-based studies. Res Aging. 2004;26(6):655–672. doi:10.1177/0164027504268574

34. Zimet GD, Dahlem NW, Zimet SG, Farley GK. The Multidimensional Scale of Perceived Social Support. J Pers Assess. 1988;52(1):30–41. doi:10.1207/s15327752jpa5201_2

35. Brinkhof LP, de Wit S, Murre JMJ, Krugers HJ, Ridderinkhof KR. The subjective experience of ageism: the Perceived Ageism Questionnaire (PAQ). Int J Environ Res Public Health. 2022;19(14):8792. doi:10.3390/ijerph19148792

36. Diener E, Emmons RA, Larsen RJ, Griffin S. The Satisfaction With Life Scale. J Pers Assess. 1985;49(1):71–75. doi:10.1207/s15327752jpa4901_13

37. Kroenke K, Spitzer RL, Williams JBW. The PHQ-9: validity of a brief depression severity measure. J Gen Intern Med. 2001;16(9):606–613. doi:10.1046/j.1525-1497.2001.016009606.x

38. Ware JE Jr, Sherbourne CD. The MOS 36-Item Short-Form Health Survey (SF-36): I. Conceptual framework and item selection. Med Care. 1992;30(6):473–483. doi:10.1097/00005650-199206000-00002

39. Emanuelsson I, Reuterberg SE, Svensson A. Changing differences in intelligence? Comparisons between groups of 13-year-olds tested from 1960 to 1990. Scand J Educ Res. 1993;37(4):259–277. doi:10.1080/0031383930370401

40. Fischer M, Heckley G, Karlsson M, Nilsson T. Revisiting Sweden’s comprehensive school reform: Effects on education and earnings. J Appl Econom. 2022;37(4):811–819. doi:10.1002/jae.2881

41. Erikson R, Rudolphi F. Change in social selection to upper secondary school—primary and secondary effects in Sweden. Eur Sociol Rev. 2010;26(3):291–305. doi:10.1093/esr/jcp022

42. Svensson A. On equality and university education in Sweden. Scand J Educ Res. 1980;24(2):79–92. doi:10.1080/0031383800240202

43. Andersson L, Allebeck P, Gustafsson JE, Gunnell D. Association of IQ scores and school achievement with suicide in a 40-year follow-up of a Swedish cohort. Acta Psychiatr Scand. 2008;118(2):99–105. doi:10.1111/j.1600-0447.2008.01171.x

44. Lundin A, Sörberg Wallin A, Falkstedt D, Allebeck P, Hemmingsson T. Intelligence and disability pension in Swedish men and women followed from childhood to late middle age. PLoS One. 2015;10(6):e0128834. doi:10.1371/journal.pone.0128834

45. MacCabe JH, Wicks S, Löfving S, David AS, Berndtsson Å, Gustafsson JE, et al. Decline in cognitive performance between ages 13 and 18 years and the risk for psychosis in adulthood: a Swedish longitudinal cohort study in males. JAMA Psychiatry. 2013;70(3):261–270. doi:10.1001/2013.jamapsychiatry.43

46. Lindenberger U, Singer T, Baltes PB. Longitudinal selectivity in aging populations: Separating mortality-associated versus experimental components in the Berlin Aging Study (BASE). J Gerontol B Psychol Sci Soc Sci. 2002;57(6):P474–P482. doi:10.1093/geronb/57.6.P474

47. Ludvigsson JF, Appelros P, Askling J, Byberg L, Carrero JJ, Ekström AM, et al. Adaptation of the Charlson comorbidity index for register-based research in Sweden. Clin Epidemiol. 2021;13:21–41. doi:10.2147/CLEP.S282475

