## Supplementary Material for "Cohort profile: The Swedish Evaluation Through Follow-up Study of Learning Later in Life (UGU-LIFE)"

**Table 1**

*Overview of data collected as part of the childhood baseline assessment in UGU at age 13.*

| Data type | Content | Note |
|---|---|---|
| Survey | Leisure activities: outdoor, verbal, technical, domestic, social, clerical | Scales developed for UGU, item level indicators vary across cohorts |
| | School: contact between children and parents, family's attitude towards higher education, pupil's anxiety in school situations, student's interest in schoolwork, student's peer contacts | Scales in UGU 1948 cohort |
| | School: family's attitude towards higher education, pupil's anxiety in school situations, pupil adjustment to school | Scales in UGU 1953 cohort |
| | Frequency of leisure activities: reading, TV, radio, cinema, sports, music, friends, homework | Single item indicators |
| | School plans and expectations, expectations from parents and teachers | Single item indicators |
| | Occupational plans, expectations from parents, friends' occupational plans | Single item indicators |
| Cognitive tests | Opposites (verbal knowledge) | Developed for UGU: 40 items, 4 options per item, 10 minutes |
| | Number series (inductive reasoning) | Developed for UGU: 40 items, 6 numbers given and 2 requested per item, 18 minutes |
| | Metal folding (spatial ability) | Developed for UGU: 40 items, 4 options per item, 15 minutes |
| School administrative data | Birth year, birth month, sex, parents' occupation and education, municipality, distance between home and school | Parental education and occupation school administrative variable in the 1948 cohort, survey question in the 1953 cohort |
| | School form, class type, class structure, class character | 1961–1969 in the 1948 cohort; 1966–1974 in the 1953 cohort |
| | Subject grades: Swedish, English, German, French, Mathematics, Physics, Latin | 1961–1969 in the 1948 cohort; 1966–1974 in the 1953 cohort |
| | Standardized knowledge tests: reading, writing, mathematics, English | In grade 6 (1961 in the 1948 cohort, 1966 in the 1953 cohort) |

**Table 2**

*Overview of data collected as part of the follow-up assessment in late adulthood (age 72/77 years).*

| Data type | Content | Note |
|---|---|---|
| Survey | Family (civil status, relationship satisfaction, children, caregiving), household (type, size), and financial (cash margin, financial satisfaction) conditions | Single item indicators |
| | Retrospective school assessment (15 items) | Developed for UGU-LIFE to match content in childhood questionnaire |
| | Leisure activities (15 items) across the lifespan: physical, reading, social | Developed for UGU-LIFE to match content in childhood questionnaire |
| | Work status, occupational history (number of jobs, extent, duration, conditions) | Single item indicators |
| | Informal training at work (3 items) | Adapted from Survey of Adult Skills (PIAAC) |
| | Informal learning engagement (18 items) and perceived competence (18 items) within theoretical (verbal, numeric, logical) and practical (mechanic, digital, motoric) knowledge domains | Developed for UGU-LIFE |
| | Internet use (8 items) | Developed for UGU-LIFE |
| | Broad Learning Adult Questionnaire (BLAQ), adapted (18 items) | Leanos, S., Coons, J., Rebok, G. W., Ozer, D. J., & Wu, R. (2019). Development of the broad learning adult questionnaire. *The International Journal of Aging and Human Development*, *88*(3), 286–311. https://doi.org/10.1177/0091415018784695 |
| | Achievement Motives Scale (AMS-R; 10 items) | Lang, J. W., & Fries, S. (2006). A revised 10-item version of the Achievement Motives Scale. *European Journal of Psychological Assessment*, *22*(3), 216–224. https://doi.org/10.1027/1015-5759.22.3.216 |
| | Basic Psychological Need Satisfaction and Frustration Scale (BPNSFS), satisfaction subscale (12 items) | Chen, B., Vansteenkiste, M., Beyers, W., Boone, L., Deci, E. L., Van der Kaap-Deeder, J., Duriez, B. Lens, W., Matos, L., Mouratidis, A., Ryan, R. M., Sheldon, K. M., Soenens, B., Van Petegem, S., & Verstuyf, J. (2015). Basic psychological need satisfaction, need frustration, and need strength across four cultures. *Motivation and Emotion*, *39*, 216–236. https://doi.org/10.1007/s11031-014-9450-1 |
| | Achievement Emotions Questionnaire, short form (AEQ-S), adapted (16 items) | Bieleke, M., Gogol, K., Goetz, T., Daniels, L., & Pekrun, R. (2021). The AEQ-S: A short version of the Achievement Emotions Questionnaire. *Contemporary Educational Psychology*, *65*, 101940. https://doi.org/10.1016/j.cedpsych.2020.101940 |
| | Implicit Theories of Intelligence (ITI) scale (8 items) | Dweck, C. S. (1999). *Self-theories: Their role in motivation, personality, and development*. Philadelphia: Psychology Press. |
| | Awareness of Age-Related Change scale, short form (AARC-10-SF) | Kaspar, R., Gabrian, M., Brothers, A., Wahl, H. W., & Diehl, M. (2019). Measuring awareness of age-related change: Development of a 10-item short form for use in large-scale surveys. *The Gerontologist*, *59*(3), e130–e140. https://doi.org/10.1093/geront/gnx213 |
| | International Personality Item Pool-Five-Factor Model scale, short form (MINI-IPIP; 20 items) | Donnellan, M.B., Oswald, F.L., Baird, B.M., & Lucas, R.E. (2006). The mini-IPIP scales: Tiny-yet-effective measures of the Big Five factors of personality. *Psychological Assessment*, *18*, 192–203. https://doi.org/10.1037/1040-3590.18.2.192 |
| | Metacognitive Awareness Inventory (MAI), short version (19 items) | Schraw, G., & Dennison, R. S. (1994). Assessing metacognitive awareness. *Contemporary Educational Psychology, 19*(4), 460–475. https://doi.org/10.1006/ceps.1994.1033 |
| | UCLA Loneliness Scale (3 items), adapted | Hughes, M. E., Waite, L. J., Hawkley, L. C., & Cacioppo, J. T. (2004). A short scale for measuring loneliness in large surveys: Results from two population-based studies. *Research on Aging*, *26*(6), 655–672. https://doi.org/10.1177/0164027504268574 |
| | Multidimensional Scale of Perceived Social Support (MPSS) scale (12 items) | Zimet, G. D., Dahlem, N. W., Zimet, S. G., & Farley, G. K. (1988). The Multidimensional Scale of Perceived Social Support. *Journal of Personality Assessment*, *52*(1), 30–41. https://doi.org/10.1207/s15327752jpa5201_2 |
| | Perceived Ageism Questionnaire (PAQ; 8 items) | Brinkhof, L. P., de Wit, S., Murre, J. M., Krugers, H. J., & Ridderinkhof, K. R. (2022). The subjective experience of ageism: the Perceived Ageism Questionnaire (PAQ). *International Journal of Environmental Research and Public Health*, *19*(14), 8792. https://doi.org/10.3390/ijerph19148792 |
| | Satisfaction With Life Scale (SWLS; 5 items) | Diener, E., Emmons, R. A., Larsen, R. J., & Griffin, S. (1985). The Satisfaction With Life Scale. *Journal of Personality Assessment*, *49*(1), 71–75. https://doi.org/10.1207/s15327752jpa4901_13 |

| Data type | Content | Note |
|---|---|---|
| | Patient Health Questionnaire (PHQ-9; 9 items) | Kroenke, K., Spitzer, R. L., & Williams, J. B. (2001). The PHQ-9: validity of a brief depression severity measure. *Journal of General Internal Medicine*, *16*(9), 606–613. https://doi.org/10.1046/j.1525-1497.2001.016009606.x |
| | Short Form Health survey instrument (SF-36; 36 items) | Ware Jr, J. E., & Sherbourne, C. D. (1992). The MOS 36-ltem short-form health survey (SF-36): I. Conceptual framework and item selection. *Medical Care*, *30*(6), 473–483. https://doi.org/10.1097/00005650-199206000-00002 |
| Cognitive tests | Opposites (verbal knowledge) | Five new words added at the end of the test to increase difficulty, otherwise identical to childhood assessment, but online |
| | Number series (inductive reasoning) | Identical to childhood assessment, but online |
| | Metal folding (spatial ability) | Identical to childhood assessment, but online |
| | Post-test confidence judgements of 10 pre-selected items from each test. | Not in childhood assessment |
| Learning tasks | Food-ordering task (procedural learning) | Developed for UGU-LIFE: 8 trials, 3 sequences (1,1,1,1,2,1,3,1), time per trial |
| | Vocabulary learning task (declarative learning) | Developed for UGU-LIFE: 10 word pairs (Sindarin–Swedish), 60 seconds training, 3 trials |
| Saliva sampling for DNA extraction | Self-administered, non-invasive DNA collection kit (Oragene, DNA Genotek) | Distributed and collected via mail. DNA extracted at Karolinska Institutet Biobank (KIBB). Genotyping performed using a genome-wide genotyping array (Illumina Infinium Global Screening Array – Multiple Disease version 3 [GSA-MD v3]). |
| Registry data from Statistics Sweden | | |
| Register of the Total Population (RTB) | Background variables: sex, year of birth, country of birth, region of birth, Swedish/foreign background, maternal and paternal birth country, migrations | |
| Population and Housing Census (FOB) | Demographics (place of residence, civil status, family, household, citizenship), education (level, orientation, examination year), occupation (status, profession, employer), income (total and disposable income, employment, childcare, housing) | Available in five-year intervals between 1960–1990 |
| Longitudinal Integrated Database for Health Insurance and Labour Market Studies (LISA) | Demographics (place of residence, civil status, family, household, citizenship), education (level and orientation (SUN), examination year), occupation (status, profession (SSYK), employer), income (total and disposable income, employment, capital, pension, disability, unemployment, parental, studies) | Available from 1990 |
| Registry data from the National Board of Health and Welfare | | |
| National Patient Register (NPR) | Inpatient and outpatient care: admission dates, caregiver, diagnoses and causes of injury (ICD codes) | Inpatient care: partial (regional) cover from 1964, full cover in psychiatric care from 1973, and national coverage from 1987. Outpatient care: partial cover from 1997, full cover from 2001. |
| National Prescribed Drug Register (PDR) | ATC code, drug name, strength, pack size, quantity, date of prescription | Available from 2005 |
| National Cause of Death Register (CDR) | Death date, cause of death (ICD codes) | Full coverage |
| Registry data from the Swedish National Archives | Conscription data (INSARK): medical examination, physical performance, psychological assessment. | From 1971 (1953 cohort only) |

**Table 3**

*Sample characteristics at baseline and follow-up.*

| | | Baseline sample | | | | | | Follow-up sample | | | | | |
|---|---|---|---|---|---|---|---|---|---|---|---|---|---|
| | | All (*N* = 21,872) | | Birth cohort 1948 (*N* = 11,945) | | Birth cohort 1953 (*N* = 9,927) | | All (*N* = 5,738) | | Birth cohort 1948 (*N* = 2,882) | | Birth cohort 1953 (*N* = 2,856) | |
| Variable | Scale range / values included | *n* (%) | *M* (*SD*) | *n* (%) | *M* (*SD*) | *n* (%) | *M* (*SD*) | *n* (%) | *M* (*SD*) | *n* (%) | *M* (*SD*) | *n* (%) | *M* (*SD*) |
| Birth cohort | | | | | | | | | | | | | |
| 1948 | | 11,945 (54.61) | | | | | | 2,882 (50.23) | | | | | |
| 1953 | | 9,927 (45.39) | | | | | | 2,856 (49.77) | | | | | |
| Sex | | | | | | | | | | | | | |
| Male | | 11,139 (50.93) | | 6,118 (51.22) | | 5,021 (50.58) | | 3,034 (52.88) | | 1,406 (48.79) | | 1,298 (45.45) | |
| Female | | 10,733 (49.07) | | 5,827 (48.78) | | 4,906 (49.42) | | 2,704 (47.12) | | 1,476 (51.21) | | 1,558 (54.55) | |
| Birth region | | | | | | | | | | | | | |
| Metropolitan | Stockholm, Skåne, Västra Götaland | 6,689 (31.54) | | 3,635 (31.28) | | 3,054 (31.85) | | 1,763 (31.19) | | 912 (32.10) | | 851 (30.27) | |
| Non-metropolitan | All other | 14,519 (68.46) | | 7,985 (68.72) | | 6,534 (68.15) | | 3,389 (68.81) | | 1,929 (67.90) | | 1,960 (69.73) | |
| Region of residence 2024 | | | | | | | | | | | | | |
| Metropolitan | Stockholm, Skåne, Västra Götaland | 7,675 (45.54) | | 3,975 (45.35) | | 3,700 (45.75) | | 2,695 (46.99) | | 1,378 (47.85) | | 1,317 (46.13) | |
| Non-metropolitan | All other | 9,178 (54.46) | | 4,790 (54.65) | | 4,388 (54.25) | | 3,040 (53.01) | | 1,502 (52.15) | | 1,538 (53.87) | |
| Marital status 1990 | | | | | | | | | | | | | |
| Married | Married | 13,539 (64.91) | | 7,645 (67.13) | | 5,894 (62.25) | | 4,107 (71.95) | | 2,126 (74.13) | | 1,981 (69.75) | |
| Unmarried | Unmarried, Divorced, Widowed | 7,318 (35.09) | | 3,743 (32.87) | | 3,575 (37.75) | | 1,601 (28.05) | | 742 (25.87) | | 859 (30.25) | |
| Marital status 2024 | | | | | | | | | | | | | |
| Married | Married, Registered partner | 9,501 (56.38) | | 4,930 (56.25) | | 4,571 (56.52) | | 3,541 (61.74) | | 1,767 (61.35) | | 1,774 (62.14) | |
| Unmarried | Unmarried, Divorced, Widowed | 7,352 (43.62) | | 3,835 (43.75) | | 3,517 (43.48) | | 2,194 (38.26) | | 1,113 (38.65) | | 1,081 (37.86) | |
| Parental background | | | | | | | | | | | | | |
| Swedish | Born in Sweden with two Swedish-born parents | 19,880 (91.26) | | 11,097 (93.03) | | 8,783 (89.13) | | 5,263 (91.74) | | 2,687 (93.23) | | 2,576 (90.23) | |
| Foreign | Foreign-born, Born in Sweden with one/two foreign-born parents | 1,903 (8,74) | | 832 (6.98) | | 1,071 (10.87) | | 474 (8.26) | | 195 (6.77) | | 279 (9.77) | |
| Educational level mother | 1–4 | 20,861 | 1.20 (0.51) | 11,854 | 1.18 (0.49) | 9,007 | 1.24 (0.54) | 5,497 | 1.26 (0.58) | 2,859 | 1.24 (0.57) | 2,638 | 1.28 (0.59) |
| | Elementary[1] / Primary[2] school (1) | 17,533 (84.05) | | 10,208 (86.11) | | 7,325 (81.33) | | 4,398 (80.01) | | 2,332 (81.57) | | 2,066 (78.32) | |
| | Secondary[1] / Junior secondary[2] school (2) | 2,576 (12.35) | | 1,267 (10.69) | | 1,309 (14.53) | | 826 (15.03) | | 396 (13.85) | | 430 (16.30) | |
| | Upper secondary school[1] / Graduated[2] or equivalent (3) | 596 (2.86) | | 291 (2.45) | | 305 (3.39) | | 213 (3.88) | | 98 (3.43) | | 115 (4.36) | |
| | Diploma[1] / Academic degree[2] (4) | 156 (0.75) | | 88 (0.74) | | 68 (0.76) | | 60 (1.09) | | 33 (1.15) | | 27 (1.02) | |
| Educational level father | 1–4 | 21,038 | 1.31 (0.75) | 11,480 | 1.28 (0.72) | 9,558 | 1.35 (0.78) | 5,561 | 1.40 (0.83) | 2,787 | 1.37 (0.82) | 2,774 | 1.42 (0.85) |

| | | Baseline sample | | | | | | Follow-up sample | | | | | |
|---|---|---|---|---|---|---|---|---|---|---|---|---|---|
| | | All ($N$ = 21,872) | | Birth cohort 1948 ($N$ = 11,945) | | Birth cohort 1953 ($N$ = 9,927) | | All ($N$ = 5,738) | | Birth cohort 1948 ($N$ = 2,882) | | Birth cohort 1953 ($N$ = 2,856) | |
| Variable | Scale range / values included | *n* (%) | *M* (*SD*) | *n* (%) | *M* (*SD*) | *n* (%) | *M* (*SD*) | *n* (%) | *M* (*SD*) | *n* (%) | *M* (*SD*) | *n* (%) | *M* (*SD*) |
| | Elementary[1] / Primary[2] school (1) | 17,331 (82.38) | | 9,724 (84.70) | | 7,607 (79.59) | | 4,345 (78.13) | | 2,227 (79.91) | | 2,118 (76.35) | |
| | Secondary[1] / Junior secondary[2] school (2) | 1,625 (84.05) | | 741 (6.46) | | 884 (9.25) | | 503 (9.05) | | 220 (7.89) | | 283 (10.20) | |
| | Upper secondary school[1] / Graduated[2] or equivalent (3) | 1,318 (6.27) | | 614 (5.35) | | 704 (7.37) | | 434 (7.80) | | 201 (7.21) | | 233 (8.40) | |
| | Diploma[1] / Academic degree[2] (4) | 764 (3.63) | | 401 (3.49) | | 363 (3.80) | | 279 (5.02) | | 139 (4.99) | | 140 (5.05) | |
| Educational level 1990 | 1–7 | 20,819 | 3.54 (1.55) | 11,362 | 3.49 (1.62) | 9,457 | 3.60 (1.46) | 5,705 | 4.13 (1.52) | 2,867 | 4.18 (1.60) | 2,838 | 4.07 (1.43) |
| | Pre-secondary education shorter than 9 years (1) | 1,928 (9.26) | | 1,556 (13.69) | | 372 (3.93) | | 230 (4.03) | | 184 (6.42) | | 46 (1.62) | |
| | Pre-secondary education shorter than 9 years (2) | 3,304 (15.87) | | 1,444 (12.71) | | 1,860 (19.67) | | 546 (9.57) | | 225 (7.85) | | 321 (11.31) | |
| | Upper secondary education maximum 2 years (3) | 6,925 (33.26) | | 3,613 (31.80) | | 3,312 (35.02) | | 1,622 (28.43) | | 721 (25.15) | | 901 (31.75) | |
| | Upper secondary education 3 years (4) | 2,530 (12.15) | | 1,481 (13.03) | | 1,049 (11.09) | | 772 (13.53) | | 421 (14.68) | | 351 (12.37) | |
| | Post-secondary education shorter than 3 years (5) | 2,703 (12.98) | | 1,302 (11.46) | | 1,401 (14.81) | | 1,012 (17.74) | | 442 (15.42) | | 570 (20.08) | |
| | Post-secondary education 3 years or longer (excl. postgraduate education) (6) | 3,294 (15.82) | | 1,881 (16.56) | | 1,413 (14.94) | | 1,456 (25.52) | | 825 (28.78) | | 631 (22.23) | |
| | Postgraduate education (7) | 135 (0.65) | | 85 (0.75) | | 50 (0.53) | | 67 (1.17) | | 49 (1.71) | | 18 (0.63) | |
| Educational level 2024 | 1–7 | 16,827 | 3.75 (1.57) | 8,749 | 3.66 (1.64) | 8,078 | 3.84 (1.49) | 5,728 | 4.28 (1.52) | 2,876 | 4.26 (1.60) | 2,852 | 4.29 (1.44) |
| | Pre-secondary education shorter than 9 years (1) | 1,280 (7.61) | | 1,027 (11.74) | | 253 (3.13) | | 207 (3.61) | | 168 (5.84) | | 39 (1.37) | |
| | Pre-secondary education shorter than 9 years (2) | 2,185 (12.99) | | 950 (10.86) | | 1,235 (15.29) | | 444 (7.75) | | 206 (7.16) | | 238 (8.35) | |
| | Upper secondary education maximum 2 years (3) | 5,397 (32.07) | | 2,708 (30.95) | | 2,689 (33.29) | | 1,522 (26.57) | | 702 (24.41) | | 820 (28.75) | |
| | Upper secondary education 3 years (4) | 2,168 (12.88) | | 1,171 (13.38) | | 997 (12.34) | | 788 (13.76) | | 417 (14.50) | | 371 (13.01) | |
| | Post-secondary education shorter than 3 years (5) | 2,487 (14.78) | | 1,142 (13.05) | | 1,345 (16.65) | | 1,058 (18.47) | | 458 (15.93) | | 600 (21.04) | |
| | Post-secondary education 3 years or longer (excl. postgraduate education) (6) | 3,075 (18.27) | | 1,640 (18.75) | | 1,435 (17.76) | | 1,576 (27.51) | | 855 (29.73) | | 721 (25.28) | |
| | Postgraduate education (7) | 235 (1.40) | | 111 (1.27) | | 124 (1.54) | | 133 (2.32) | | 70 (2.43) | | 63 (2.21) | |
| General cognitive ability[3] age 13 | 3–116 | 19,928 | 64.18 (18.11) | 10,556 | 62.69 (18.07) | 9,372 | 65.85 (18.00) | 5,352 | 70.29 (17.03) | 2,612 | 69.09 (17.19) | 2,740 | 71.44 (16.79) |
| Disposable income (in 100 SEK) 1990 | 0–74,725 | 20,857 | 1,189.76 (727.81) | 11,388 | 1,211.62 (887.56) | 9,469 | 1,163.47 (467.08) | 5,708 | 1,272.79 (551.24) | 2,868 | 1,337.07 (627.21) | 2,840 | 1,207.87 (452.96) |
| Disposable income (in 100 SEK) 2023 | -224,396–1,160,362 | 17,152 | 3,245.57 (10,314.56) | 8,970 | 2,921.36 (4,204.60) | 8,182 | 3,601.00 (14,262.46) | 5,733 | 3,558.57 (4,555.70) | 2,880 | 3,380.26 (4,325.31) | 2,853 | 3,738.57 (4,771.03) |
| Charlson Comorbidity Index[4] 1990 | 0–7 | 10,376 | 0.09 (0.44) | 5,174 | 0.10 (0.47) | 5,202 | 0.07 (0.41) | 3,354 | 0.04 (0.27) | 1,598 | 0.04 (0.29) | 1,756 | 0.04 (0.24) |
| Charlson Comorbidity Index[4] 2025 | 0–18 | 19,602 | 1.45 (2.11) | 10,828 | 1.67 (2.24) | 8,774 | 1.18 (1.92) | 5,207 | 0.80 (1.38) | 2,638 | 0.89 (1.43) | 3,569 | 0.71 (1.33) |

*Note*. [1]1948 cohort, administrative data. [2]1953 cohort, survey question. [3]Sum score of verbal knowledge (opposites), inductive reasoning (number series), and metal folding (spatial ability) tests. [4]Weighted score calculated with code provided by Ludvigsson et al., 2021 (https://github.com/bjoroeKI/Charlson-comorbidity-index-revisited).

**Table 4**

*Missing data patterns across variables and data sources.*

| Variable | Source | Missing: All (*N* = 21,872) *n* | % | Birth cohort 1948 (*N* = 11,945) *n* | % | Birth cohort 1953 (*N* = 9,927) *n* | % | % of missing identified as deceased in CDR |
|---|---|---|---|---|---|---|---|---|
| Birth cohort | UGU 1961/1966 | 0 | | 0 | | 0 | | |
| Sex | UGU 1961/1966 | 0 | | 0 | | 0 | | |
| Birth region | RTB | 664 | 3.04 | 325 | 2.72 | 339 | 3.41 | 22.89 |
| Region of residence 2024 (age 71/76) | LISA 2024 | 5,019 | 22.94 | 3,180 | 26.62 | 1,839 | 18.53 | 87.67 |
| Marital status 1990 (age 37/42) | LISA 1990 | 1,014 | 4.64 | 557 | 4.66 | 458 | 4.61 | 47.49 |
| Marital status 2024 (age 71/76) | LISA 2024 | 5,019 | 22.95 | 3,180 | 26.62 | 1,839 | 18.53 | 87.67 |
| Parental background | RTB | 89 | 0.41 | 16 | 0.13 | 73 | 0.74 | 0 |
| Educational level mother | UGU 1961/1966 | 1,011 | 4.62 | 91 | 0.76 | 920 | 9.27 | 19.78 |
| Educational level father | UGU 1961/1966 | 834 | 3.81 | 465 | 3.89 | 369 | 3.72 | 26.86 |
| Educational level 1990 (age 37/42) | LISA 1990 | 1,053 | 4.81 | 583 | 4.88 | 470 | 4.73 | 47.39 |
| Educational level 2024 (age 71/76) | LISA 2024 | 5,045 | 23.07 | 3,196 | 26.76 | 1,849 | 18.63 | 87.22 |
| General cognitive ability age 13 (1961/1966) | UGU 1961/1966 | 1,944 | 8.89 | 1,389 | 11.63 | 555 | 5.59 | 26.29 |
| Disposable income (in 100 SEK) 1990 (age 37/42) | LISA 1990 | 1,015 | 4.64 | 557 | 4.66 | 458 | 4.61 | 47.49 |
| Disposable income (in 100 SEK) 2023 (age 70/75) | LISA 2023 | 4,720 | 21.58 | 2,975 | 24.91 | 1,745 | 17.58 | 86.74 |
| Charlson Comorbidity Index 1990 (age 37/42) | NPR 1964*–1990 | 11,496 | 52.56 | 6,771 | 56.68 | 4,725 | 47.60 | 31.23 |
| Charlson Comorbidity Index 2025 (age 72/77) | NPR 1964*–2025 | 2,270 | 10.38 | 1,117 | 9.35 | 1,153 | 11.61 | 14.63 |

*Note*. Data screened for reuse of personal identity numbers (in RTB); observations in LISA and NPR set to missing for 3 individuals with data recorded after death date (in CDR). UGU = Utvärdering Genom Uppföljning (Evaluation Through Follow-up). RTB = Register of the Total Population. LISA = Longitudinal Integrated Database for Health Insurance and Labour Market Studies. NPR = National Patient Register, inpatient care. CDR = Cause of Death Registry (by 2025-10-26). *National coverage from 1987.

**Table 5**

*Standardized mean differences (SD) between the baseline sample and the sample at follow-up, decomposed into mortality-associated, experimental, and total selectivity.*

| | All | | | Birth cohort 1948 | | | Birth cohort 1953 | | |
|---|---|---|---|---|---|---|---|---|---|
| Variable | Survival | Experimental | Total | Survival | Experimental | Total | Survival | Experimental | Total |
| Birth cohort (1953 vs. 1948) | 0.060 | 0.034 | 0.088 | | | | | | |
| Sex (male vs. female) | -0.054 | -0.028 | -0.076 | -0.057 | 0.002 | -0.049 | -0.050 | -0.058 | -0.103 |
| Birth region (metropolitan vs. non-metropolitan) | -0.006 | 0.006 | -0.007 | -0.004 | 0.030 | 0.018 | -0.010 | -0.018 | -0.034 |
| Region of residence 2024 (metropolitan vs. non-metropolitan) | -0.002 | 0.031 | 0.029 | -0.002 | 0.051 | 0.050 | -0.002 | 0.010 | 0.008 |
| Parental background (Swedish vs. foreign) | -0.004 | 0.001 | 0.017 | 0.000 | -0.012 | 0.008 | -0.001 | 0.016 | 0.035 |
| Educational level mother | 0.023 | 0.103 | 0.112 | 0.031 | 0.114 | 0.129 | 0.008 | 0.087 | 0.083 |
| Educational level father | 0.029 | 0.103 | 0.115 | 0.033 | 0.119 | 0.134 | 0.020 | 0.084 | 0.087 |
| Cognitive ability age 13 | 0.054 | 0.294 | 0.338 | 0.057 | 0.308 | 0.354 | 0.041 | 0.278 | 0.310 |
| Educational level 1990 | 0.060 | 0.325 | 0.379 | 0.068 | 0.366 | 0.428 | 0.047 | 0.282 | 0.323 |
| Educational level 2024 | 0.004 | 0.335 | 0.337 | 0.005 | 0.364 | 0.366 | 0.003 | 0.302 | 0.303 |
| Marital status 1990 (married vs. unmarried) | 0.066 | 0.083 | 0.147 | 0.079 | 0.072 | 0.149 | 0.058 | 0.099 | 0.155 |
| Marital status 2024 (married vs. unmarried) | 0.005 | 0.106 | 0.108 | 0.005 | 0.101 | 0.103 | 0.004 | 0.111 | 0.113 |
| Disposable income 1990 | 0.022 | 0.090 | 0.114 | 0.027 | 0.112 | 0.141 | 0.021 | 0.074 | 0.095 |
| Disposable income 2023 | 0.002 | 0.029 | 0.030 | 0.003 | 0.107 | 0.109 | 0.001 | 0.009 | 0.010 |
| Charlson Comorbidity Index 1990 | -0.089 | -0.013 | -0.100 | -0.103 | -0.019 | -0.120 | -0.073 | -0.006 | -0.078 |
| Charlson Comorbidity Index 2025 | -0.251 | -0.071 | -0.306 | -0.277 | -0.086 | -0.346 | -0.210 | -0.049 | -0.246 |

**Table 6**

*Frequency of task completion at follow-up assessment in late adulthood.*

| Task | Data preparation note* | All (*N* = 5,738) | | Birth cohort 1948 (*N* = 2,882) | | Birth cohort 1953 (*N* = 2,856) | |
|---|---|---|---|---|---|---|---|
| | | *n* | % | *n* | % | *n* | % |
| Food-ordering task | 65 observations 100% missing values; 2 observations after death date | 2,711 | | 1,300 | | 1,411 | |
| Vocabulary learning task | 200 observations missing in trial 1; 2 observations after death date | 2,564 | | 1,225 | | 1,339 | |
| Opposites | 64 observations >85% missing values; 1 observation missing in 20 first items; 1 observation after death date | 2,647 | | 1,267 | | 1,380 | |
| Number series | 6 duplicate observations; 197 observations >86% missing values | 2,175 | | 1,023 | | 1,152 | |
| Metal folding | 116 observations >85% missing values; 1 observation missing in 20 first items | 2,369 | | 1,134 | | 1,235 | |
| Survey | 184 duplicate observations; 29 observations 100% missing values; 7 observations >98% missing values; 1 observation after death date | 5,363 | | 2,712 | | 2,651 | |
| Online | | 2,207 | 41.15 | 1,033 | 38.09 | 1,174 | 44.29 |
| Paper | | 3,156 | 58.85 | 1,679 | 61.91 | 1,477 | 55.71 |
| Saliva for DNA extraction | Samples collected between the 27th of June 2025 and the 12th of May 2026 | 2,166 | | 1,078 | | 1,088 | |
| Number of tasks completed, *M* (*SD*) | | 3.48 (2.50) | | 3.38 (2.45) | | 3.59 (2.54) | |
| 1 | | 2,217 | 38.64 | 1,123 | 38.97 | 1,094 | 38.31 |
| 2 | | 809 | 14.10 | 450 | 15.61 | 359 | 12.57 |
| 3 | | 184 | 3.21 | 95 | 3.30 | 89 | 3.12 |
| 4 | | 171 | 2.98 | 86 | 2.98 | 85 | 2.98 |
| 5 | | 303 | 5.28 | 171 | 5.93 | 132 | 4.62 |
| 6 | | 969 | 16.89 | 467 | 16.20 | 502 | 17.58 |
| 7 | | 1,085 | 18.91 | 490 | 17.00 | 595 | 20.83 |

*Note*. *Observations removed from original (raw) data files as part of data cleaning. Data screened for duplicate responses, missing values, and data flagged as potentially invalid due to recorded death date (Cause of Death Registry) and reuse of personal identity numbers (Register of the Total Population).

**Table 7**

*Standardized mean differences (SD) between each task-specific sample and the total follow-up sample.*

| Variable | Food-ordering task | Vocabulary learning task | Opposites | Number series | Metal folding | Online survey | Paper survey | Saliva for DNA extraction |
|---|---|---|---|---|---|---|---|---|
| Birth cohort (1953 vs. 1948) | 0.045 | 0.049 | 0.047 | 0.064 | 0.047 | 0.068 | -0.059 | 0.009 |
| Sex (male vs. female) | | | | | | | | |
| All | -0.034 | -0.038 | -0.040 | -0.047 | -0.040 | -0.046 | 0.009 | 0.034 |
| 1948 | 0.010 | 0.012 | 0.011 | -0.004 | 0.010 | 0.010 | -0.028 | 0.089 |
| 1953 | -0.072 | -0.082 | -0.083 | -0.081 | -0.083 | -0.092 | 0.046 | -0.019 |
| Region of residence 2024 (metropolitan vs. non-metropolitan) | | | | | | | | |
| All | 0.062 | 0.049 | 0.064 | 0.066 | 0.062 | 0.063 | -0.043 | 0.032 |
| 1948 | 0.059 | 0.046 | 0.054 | 0.053 | 0.046 | 0.041 | -0.031 | -0.012 |
| 1953 | 0.066 | 0.053 | 0.075 | 0.079 | 0.078 | 0.084 | -0.060 | 0.076 |
| General cognitive ability age 13 | | | | | | | | |
| All | 0.288 | 0.325 | 0.305 | 0.367 | 0.338 | 0.341 | -0.243 | 0.199 |
| 1948 | 0.304 | 0.350 | 0.321 | 0.390 | 0.352 | 0.375 | -0.233 | 0.208 |
| 1953 | 0.269 | 0.297 | 0.286 | 0.342 | 0.321 | 0.306 | -0.246 | 0.190 |
| Educational level 2024 | | | | | | | | |
| All | 0.264 | 0.290 | 0.275 | 0.323 | 0.304 | 0.308 | -0.216 | 0.198 |
| 1948 | 0.302 | 0.319 | 0.309 | 0.364 | 0.350 | 0.338 | -0.206 | 0.213 |
| 1953 | 0.227 | 0.262 | 0.241 | 0.285 | 0.259 | 0.280 | -0.227 | 0.183 |
| Marital status 2024 (married vs. unmarried) | | | | | | | | |
| All | 0.032 | 0.039 | 0.038 | 0.050 | 0.049 | 0.042 | -0.031 | 0.039 |
| 1948 | 0.030 | 0.046 | 0.039 | 0.062 | 0.056 | 0.051 | -0.030 | 0.065 |
| 1953 | 0.034 | 0.032 | 0.037 | 0.038 | 0.041 | 0.033 | -0.031 | 0.013 |

| Variable | Food-ordering task | Vocabulary learning task | Opposites | Number series | Metal folding | Online survey | Paper survey | Saliva for DNA extraction |
|---|---|---|---|---|---|---|---|---|
| Disposable income 2023 | | | | | | | | |
| All | 0.069 | 0.084 | 0.078 | 0.096 | 0.083 | 0.090 | -0.063 | 0.041 |
| 1948 | 0.086 | 0.115 | 0.099 | 0.133 | 0.109 | 0.123 | -0.075 | 0.040 |
| 1953 | 0.051 | 0.055 | 0.057 | 0.062 | 0.057 | 0.058 | -0.046 | 0.041 |
| Charlson Comorbidity Index 2025 | | | | | | | | |
| All | -0.030 | -0.045 | -0.030 | -0.044 | -0.042 | -0.038 | 0.023 | -0.038 |
| 1948 | -0.020 | -0.019 | -0.007 | -0.019 | -0.026 | -0.008 | 0.007 | -0.030 |
| 1953 | -0.033 | -0.064 | -0.046 | -0.059 | -0.050 | -0.056 | 0.034 | -0.044 |
